# “I wish I could work on school stuff.” Investigating the Impact of Remote Learning on Undergraduate Students’ Academic Success and Mental Health during the COVID-19 Lockdown

**DOI:** 10.1101/2024.12.30.24319774

**Authors:** Joseph P. Nano, William A. Catterall, Michael L. Chang, Mina H. Ghaly

**Affiliations:** The Dartmouth Institute for Health Policy & Clinical Practice, Dartmouth Geisel School of Medicine, Hanover, NH, USA; Harvard T.H. Chan School of Public Health, Department of Environmental Health, Boston, MA, USA

**Keywords:** COVID-19, Remote Learning, Mental Health, Academic performance, University students

## Abstract

**Objective:** To investigate how remote learning has affected undergraduate students’ learning abilities, academic success, and mental health during the COVID-19 pandemic.

**Methods:** This cross-sectional study took place between April and June 2020 in the United States. Participants completed a survey consisting of demographic questions, Depression, Anxiety, and Stress Scale-21 Items (DASS-21), and an open-ended question. We used a logistic regression model on objective variables and conducted a systematic thematic analysis of the open-ended response.

**Results:** Our final sample consisted of 1,173 full-time undergraduate students in the United States. Most participants were public university students (n = 835, 71%) and reported that moving to remote learning had a negative impact on their school performance (n=802, 68%). Positive experiences in remote learning were associated with moving to their family’s house (p<0.05), living on campus in a dorm (p<0.05), and no internet issues during classes (p<0.0001). From the thematic analysis, we found six common themes among those who reported having a negative experience with remote learning: (1) adjusting to school, (2) dealing with mental health difficulties, (3) lack of motivation to do work, (4) adjusting to home environment, (5) feeling uncertain about occupational opportunities, and (6) disagreement in political views.

**Limitations:** The results of this study may not be generalizable to undergraduate students outside of the United States due to differences in lockdown restrictions.

**Conclusion:** Remote learning during COVID-19 had a negative impact on the majority of undergraduate students’ academic performance and mental health.

**Highlights:**

❖ The majority of undergraduate students reported that remote learning negatively impacted their school performance.
❖ Students with non-STEM college majors experienced higher levels of stress, anxiety, and depression compared to students with STEM college majors.
❖ We found six common themes related to negative experiences with remote learning.

## Introduction

According to the World Health Organization (WHO) and the Centers for Disease Control and Prevention (CDC), mental health illness, such as anxiety and depressive disorders, is a global health priority and public health issue in the United States (U.S.) and globally.^1–3^ The mental health of university students in particular represents a public health priority.^4,5^ Studies have shown that university students report higher levels of depression and suicidality compared to the general population, with suicide rates continuing to rise over the years.^6,7^ Teens and young adults are particularly vulnerable due to the transition from high school to college, a period notably associated with an onset of mental health problems.^8^

Depression, which encompasses multiple different depressive disorders, may start during adolescence and continue into adulthood.^9,10^ There are documented gender disparities in depression, as women are more likely than men to experience depression.^9,11^ Furthermore, depression continues to be a recurrent problem among undergraduate students with a prevalence as high as 49% on college campuses.^12,13^ Mental health illness can affect students’ energy level, concentration, and school performance.

The WHO declared the coronavirus disease outbreak of SARS-CoV-2 (COVID-19) a global pandemic on March 11, 2020.^14^ The nationwide COVID-19 lockdowns forced college students to abruptly leave their campuses, disrupting their routines and intensifying mental health challenges.^15^ This sudden transition, combined with the isolation and uncertainty of the pandemic, exacerbated existing issues, highlighting the urgent need to prioritize students’ mental well-being during this unprecedented time.

In response to the mandated lockdowns, many academic institutions transitioned from in-person learning to remote learning.^16^ As such, this led to subsequent changes to class syllabi and increased workload on educators.^16^ There are differences between a typical planned online course and remote-learning courses hastily created during a pandemic.^17^ There is limited research on the impact of emergency remote education on undergraduate students’ academic performance during public health crises.^17^

The purpose of this study is to investigate the impact of universities moving classes to remote learning on undergraduate students’ academic performance and mental health. We contribute to the literature both quantitatively and qualitatively by examining three key effects of the COVID-19 lockdown: first, how students adjusted their study habits; second, their overall satisfaction during the educational crisis; and third, provide recommendations for improving the remote learning experience based on reported positive and negative experiences.

## Materials and Methods

### Participants

We recruited undergraduate students enrolled in a private university, public university, or community college in the United States. For eligibility, participants were required to be at least 18 years old, enrolled as full-time undergraduate students, and members of the following graduating class years: Class of 2020, Class of 2021, Class of 2022, or Class of 2023. We conducted our recruitment phase between April and June 2020 through two methods: emails sent to undergraduate students at Boston College and online via Reddit, a popular social platform and forum. The recruitment process involved posting study invitations on over 20 university-related and survey-focused subreddits (online communities organized around specific topics), including r/SampleSize. We were unable to conduct in-person interviews due to the U.S. lockdown and public health restrictions.

### Procedure

Our survey consisted of four sections: social demographics, school adjustments, DASS-21 questions, and an optional open-ended question. The survey took approximately 10 minutes to complete. All participants provided informed consent before completing the survey, fully aware of potential risks and benefits, the confidentiality of their responses, and their right to withdraw from participation at any time. All respondents received the option to enter into a raffle to win one of 15 $10 Amazon gift cards. Our study survey was available online for nine weeks, from April to June 2020. Our study received institutional review board (IRB) approval with exemption from the Boston College IRB in April 2020, ensuring compliance with U.S. federal regulations for the protection of human subjects.

### Measures

*DASS (Depression, Anxiety, and Stress Scale) 21* Our survey included the widely used and validated 21-item DASS-21 scale to evaluate stress, anxiety, and depression levels during the COVID-19 pandemic.^18^ We categorized scores into five categories: “normal” (depression: 0–9, anxiety: 0–7, stress: 0–14), “mild” (10–13, 8–9, 15–18), “moderate” (14–20, 10–14, 19–25), “severe” (21–27, 15–19, 26–33), and “extremely severe” (28+, 20+, 34+). Participants rated their experiences on a scale of “once a week or less” (score = 0), “2–3 times a week” (= 1), “4–6 times a week” (= 2), and “7 times a week or more” (= 3). We calculated the total score to determine participants’ severity levels.

### Statistical analysis

Statistical analyses were performed using R-Studio statistical software (Version 2023.06.1+524, R-Studio, PBC). Our first phase of analysis consisted of descriptive statistics that summarized the sample’s demographics and the distribution of the three mental health outcomes (stress, anxiety, and depression) among undergraduate students. We used t-tests to examine whether the transition to remote learning impacted students’ DASS-21 scores for stress, anxiety, and depression levels. We used a predictive model to evaluate the influence of factors such as gender, frequency of leaving home, and academic performance after transitioning to remote learning. We conducted a thematic analysis of students’ responses to the open-ended question in our survey. Two independent researchers (JN and MG) read responses and captured key concepts. Using an inductive approach, reviewers applied codes to each response to extract meaning from the text. These codes were grouped into broader categories, which were then synthesized into overarching themes. All conflicts were resolved.

## Results

### Descriptive statistics

A total of 1,734 responses were received, but after excluding students under 18 (n = 34), part-time students (n = 135), and incomplete responses (n = 392), the final sample consisted of 1,173 participants. Among participants who reported negative experiences after moving to remote learning (n = 802, 68%), the majority of participants attended public universities (n = 541, 68%), followed by private universities (n = 254, 31%) (**Table 1**). For graduating class, participants were distributed across the class of 2020 (n = 130, 16%), 2021 (n = 209, 26%), 2022 (n = 227, 28%), and 2023 (n = 205, 26%). The sample’s participants identified as 47% male, 49% female, and 4% as other. Housing status varied, with nearly half of the participants reporting that they had moved back to their family’s house during the pandemic (n = 379, 47%), while a quarter of participants continued living with their parents (n = 210, 26%) (**Table 1**). Regarding internet connectivity, approximately half of participants reported no challenges (n = 421, 53%) (**Table 1**). For learning styles, the majority of participants reported preference for in-person learning (n = 716, 89%). For demographics, the majority of our participants identified as “Caucasian” (n = 401, 55%) or Asian or Pacific Islanders (n = 173, 24%). The majority of undergraduate students in this study used Zoom as their remote learning platform (n = 585, 73%), followed by “other” platforms (n = 208, 26%) (**Table 1**). We identified significant differences between students who reported a positive impact of remote learning on their academic performance and those who reported a negative impact. These differences were associated with challenges in internet connection (p<0.001), preference for type of learning (p<0.001), remote learning program used (p<0.001), and gender (p<0.05) (**Table 1**).

**Table 1.**
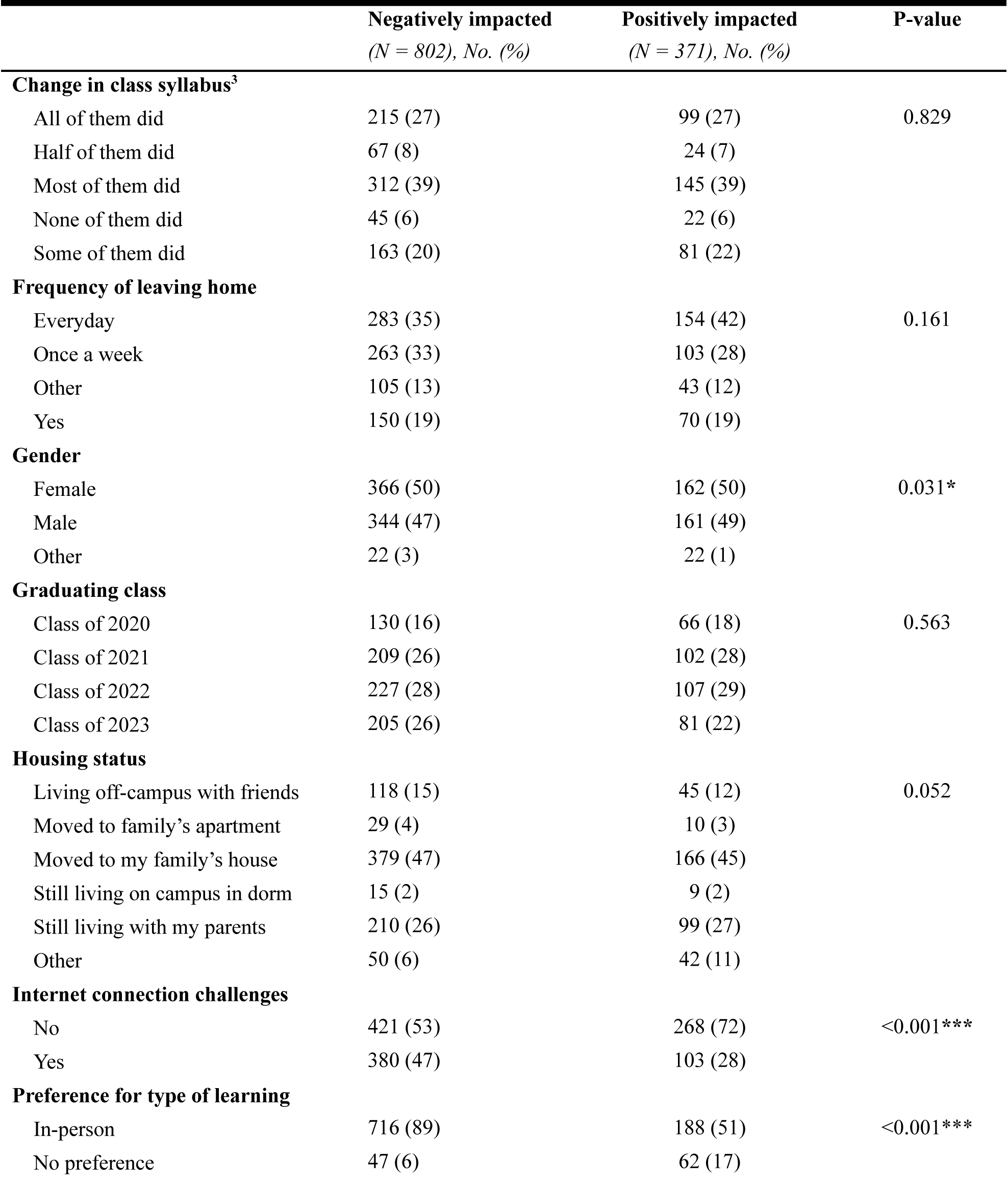
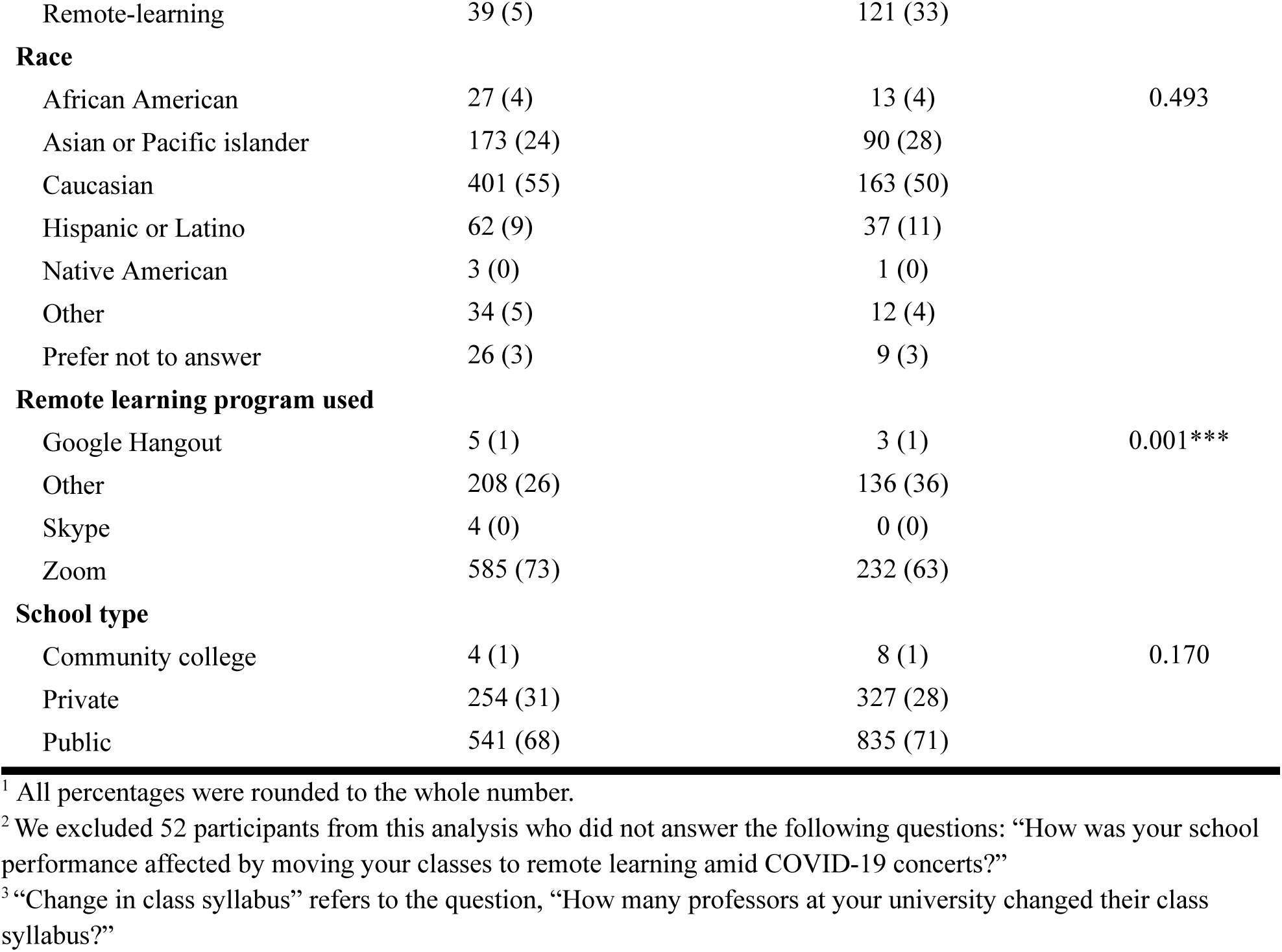
Baseline characteristics among students who stated that moving to remote learning had a negative impact on their school performance compared to students who claimed that remote learning had a positive impact on their school performance ^1,2^.

### DASS-21

Among all participants, 13% (n = 156) reported experiencing “severe” or “extremely severe” stress (**Figure 1A**). Of these participants, 57% (n = 89) were female, and 51% (n = 80) identified as Caucasian. Notably, 81% (n = 126) of participants within this group indicated that transitioning to remote learning negatively impacted their academic performance (**Figure 1A**). Furthermore, within this group, 18% (n = 28) were members of the class of 2020, and 71% (n = 111) attended public schools (**Figure 1A**).

**Figure 1.**
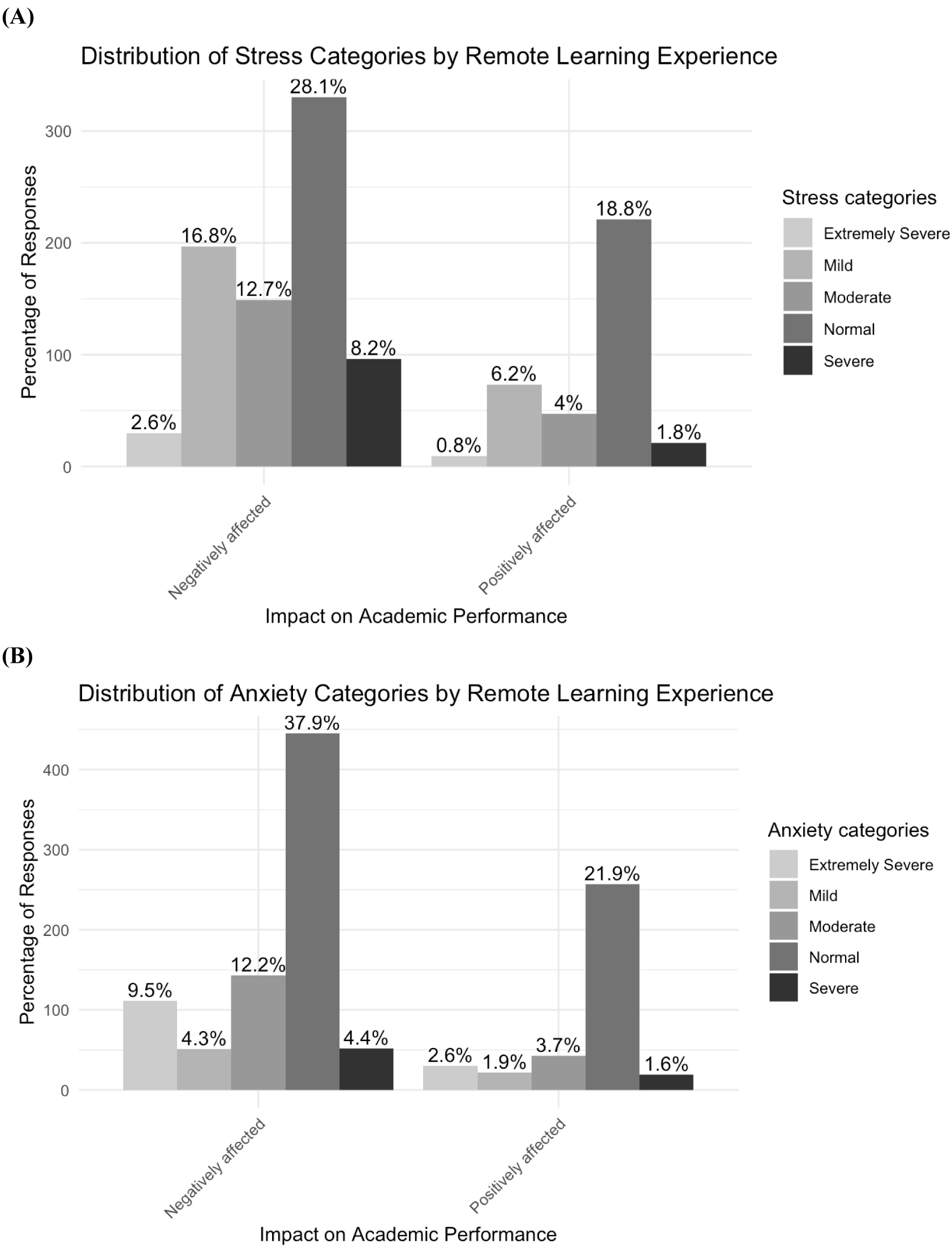
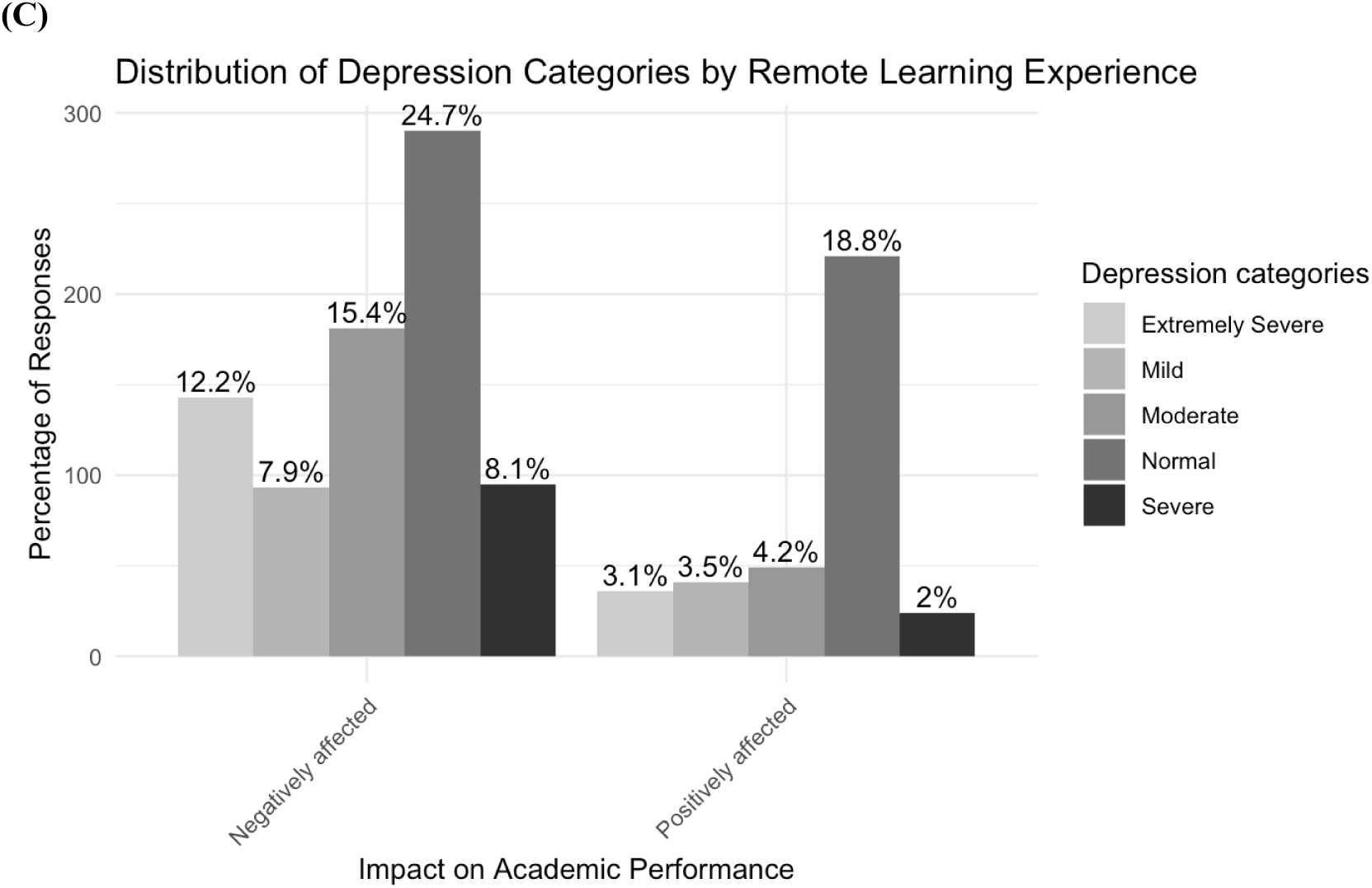
Distribution of DASS-21 categories (A) stress, (B) anxiety, and (C) depression among students who reported that moving to remote learning had a positive impact or negative impact on their school performance during the COVID-19 lockdown in the United States

Approximately 18% (n = 212) of all participants reported experiencing “severe” or “extremely severe” anxiety (**Figure 1B**). Of these participants, 56% (n = 119) were female, and 51% (n = 109) identified as Caucasian. Additionally, 77% (n = 163) of participants within this group stated that moving to remote learning negatively impacted their academic performance, (**Figure 1B**). Within this group, 16% (n = 34) were members of the class of 2020, and 71% (n = 151) attended public schools (**Figure 1B**).

Approximately 25% (n = 298) of all claimed experiencing “severe” or “extremely severe” depression (**Figure 1C**). Of these participants, 53% (n = 157) were female, and 51% (n = 153) identified as Caucasian. Furthermore, 80% (n = 238) of participants within this group reported that moving to remote learning negatively impacted their academic performance (**Figure 1C**). Within this group, 18% (n = 53) were members of the class of 2020, while 72% (n = 215) attended public schools (**Figure 1C**).

We received responses from 647 undergraduate students enrolled full-time in an educational institution with Science, Technology, Engineering, and Mathematics (STEM) majors and 520 undergraduate students enrolled full-time with non-STEM majors (**Figure 2**). We found undergraduate students with non-STEM majors to have significantly higher levels of stress (p < 0.001), anxiety (p < 0.001), and depression (p < 0.01) compared to students with STEM majors (**Figure 2**).

**Figure 2.**
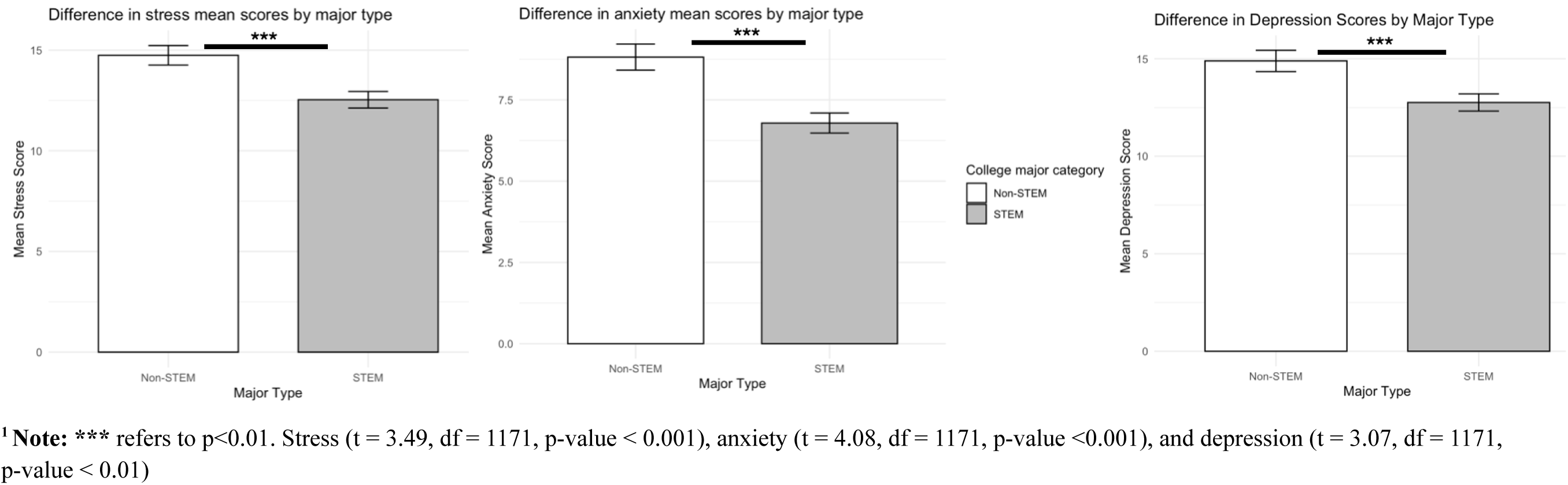
Stress, Anxiety, and Depression mean scores among undergraduate students based on college major, categorized as STEM (n = 647) and Non-STEM (n = 520) majors^1^

### Predictive model

Our logistic regression model revealed several factors significantly associated with undergraduate students’ experience with remote learning in the United States (**Table 2**). Undergraduate students who preferred remote learning (OR = 13.106, 95% CI: 8.296–20.707, p < 0.001) or had no specific preference for learning type (OR = 5.297, 95% CI: 3.267–8.590, p < 0.001) were significantly more likely to report a better remote learning experience compared to those who preferred in-person learning (**Table 2**). Regarding housing status, students who moved to their family’s house (OR = 1.939, 95% CI: 1.145–3.285, p = 0.014), stayed on campus in a dorm (OR = 3.158, 95% CI: 1.105–9.022, p = 0.032), or reported "other" housing arrangements (OR = 2.589, 95% CI: 1.275–5.253, p = 0.008) were more likely to have a different experience compared to those living off-campus (**Table 2**). Students with internet issues were significantly less likely to report a positive remote learning experience (OR = 0.448, 95% CI: 0.323–0.622, p < 0.001). Those using Zoom as a remote learning platform were less likely to have a positive experience compared to other platforms (OR = 0.610, 95% CI: 0.432–0.861, p = 0.005). No significant associations were found for gender, school type, or the number of professors who changed their syllabus (**Table 2**).

**Table 2.**
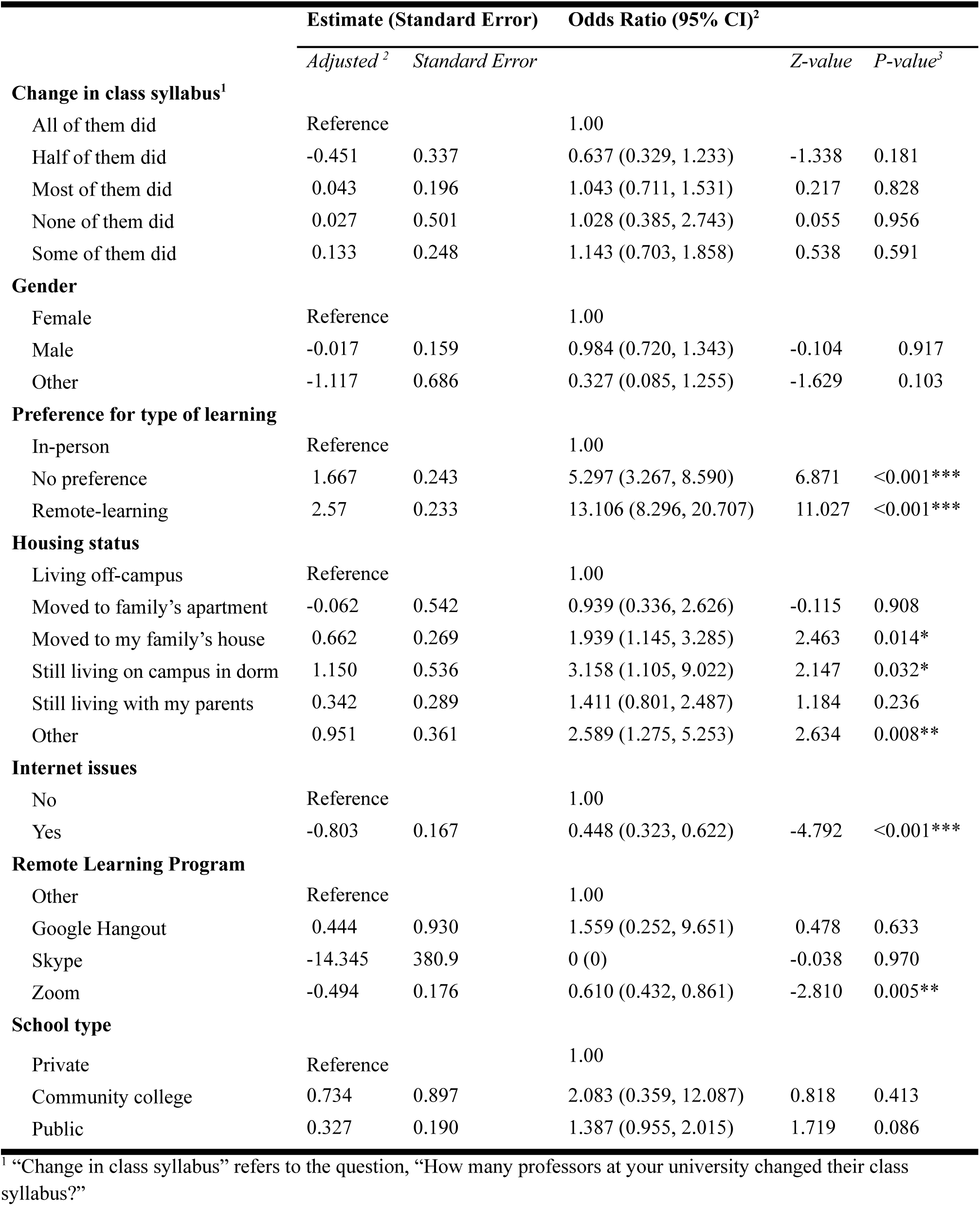
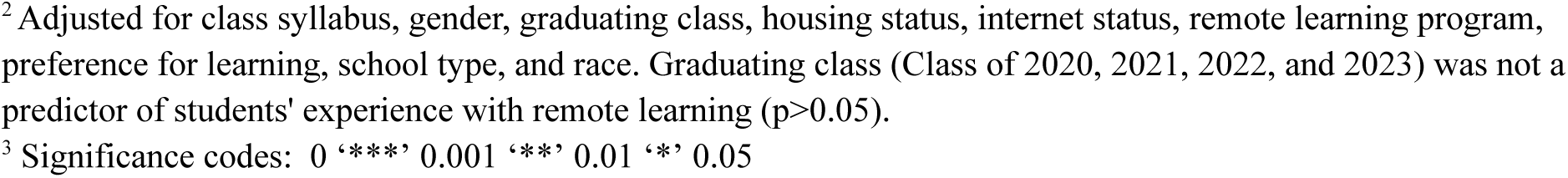
Logistic regression model analysis of sociodemographic and other factors impacting undergraduate students’ experience with remote learning experience in the United States.

|  | Estimate (Standard Error) |  | Odds Ratio (95% CI)<sup>2</sup> |  |  |
| --- | --- | --- | --- | --- | --- |
|  | *Adjusted <sup>2</sup>* | *Standard Error* |  | *Z-value* | *P-value<sup>3</sup>* |
| **Change in class syllabus<sup>1</sup>** |  |  |  |  |  |
| All of them did | Reference |  | 1.00 |  |  |
| Half of them did | -0.451 | 0.337 | 0.637 (0.329, 1.233) | -1.338 | 0.181 |
| Most of them did | 0.043 | 0.196 | 1.043 (0.711, 1.531) | 0.217 | 0.828 |
| None of them did | 0.027 | 0.501 | 1.028 (0.385, 2.743) | 0.055 | 0.956 |
| Some of them did | 0.133 | 0.248 | 1.143 (0.703, 1.858) | 0.538 | 0.591 |
| **Gender** |  |  |  |  |  |
| Female | Reference |  | 1.00 |  |  |
| Male | -0.017 | 0.159 | 0.984 (0.720, 1.343) | -0.104 | 0.917 |
| Other | -1.117 | 0.686 | 0.327 (0.085, 1.255) | -1.629 | 0.103 |
| **Preference for type of learning** |  |  |  |  |  |
| In-person | Reference |  | 1.00 |  |  |
| No preference | 1.667 | 0.243 | 5.297 (3.267, 8.590) | 6.871 | <0.001*** |
| Remote-learning | 2.57 | 0.233 | 13.106 (8.296, 20.707) | 11.027 | <0.001*** |
| **Housing status** |  |  |  |  |  |
| Living off-campus | Reference |  | 1.00 |  |  |
| Moved to family’s apartment | -0.062 | 0.542 | 0.939 (0.336, 2.626) | -0.115 | 0.908 |
| Moved to my family’s house | 0.662 | 0.269 | 1.939 (1.145, 3.285) | 2.463 | 0.014* |
| Still living on campus in dorm | 1.150 | 0.536 | 3.158 (1.105, 9.022) | 2.147 | 0.032* |
| Still living with my parents | 0.342 | 0.289 | 1.411 (0.801, 2.487) | 1.184 | 0.236 |
| Other | 0.951 | 0.361 | 2.589 (1.275, 5.253) | 2.634 | 0.008** |
| **Internet issues** |  |  |  |  |  |
| No | Reference |  | 1.00 |  |  |
| Yes | -0.803 | 0.167 | 0.448 (0.323, 0.622) | -4.792 | <0.001*** |
| **Remote Learning Program** |  |  |  |  |  |
| Other | Reference |  | 1.00 |  |  |
| Google Hangout | 0.444 | 0.930 | 1.559 (0.252, 9.651) | 0.478 | 0.633 |
| Skype | -14.345 | 380.9 | 0 (0) | -0.038 | 0.970 |
| Zoom | -0.494 | 0.176 | 0.610 (0.432, 0.861) | -2.810 | 0.005** |
| **School type** |  |  |  |  |  |
| Private | Reference |  | 1.00 |  |  |
| Community college | 0.734 | 0.897 | 2.083 (0.359, 12.087) | 0.818 | 0.413 |
| Public | 0.327 | 0.190 | 1.387 (0.955, 2.015) | 1.719 | 0.086 |

<sup>1</sup> "Change in class syllabus" refers to the question, "How many professors at your university changed their class syllabus?"
<sup>2</sup> Adjusted for class syllabus, gender, graduating class, housing status, internet status, remote learning program, preference for learning, school type, and race. Graduating class (Class of 2020, 2021, 2022, and 2023) was not a predictor of students' experience with remote learning ( $p>0.05$ ).
<sup>3</sup> Significance codes: 0 '\*\*\*' 0.001 '\*\*' 0.01 '\*' 0.05

### Thematic Analysis

We received 563 survey responses to the open-ended question, from which 215 responses contributed to the thematic analysis (**Table 3**). A majority of the responses were between two and three sentences in length. Responses that were nondescript, too brief, or lacked an otherwise unclear theme(s) were excluded. Through the thematic analysis, we identified six common themes among those who reported having a negative experience with remote learning (**Table 3**). These included: (1) adjusting to an online learning environment, (2) dealing with mental health difficulties, (3) lack of motivation to do work, (4) adjusting to the home environment, (5) feeling uncertain about future opportunities (educational, and for soon-to-be graduates, professional), and (6) political tension (**Table 3**).

**Table 3.**
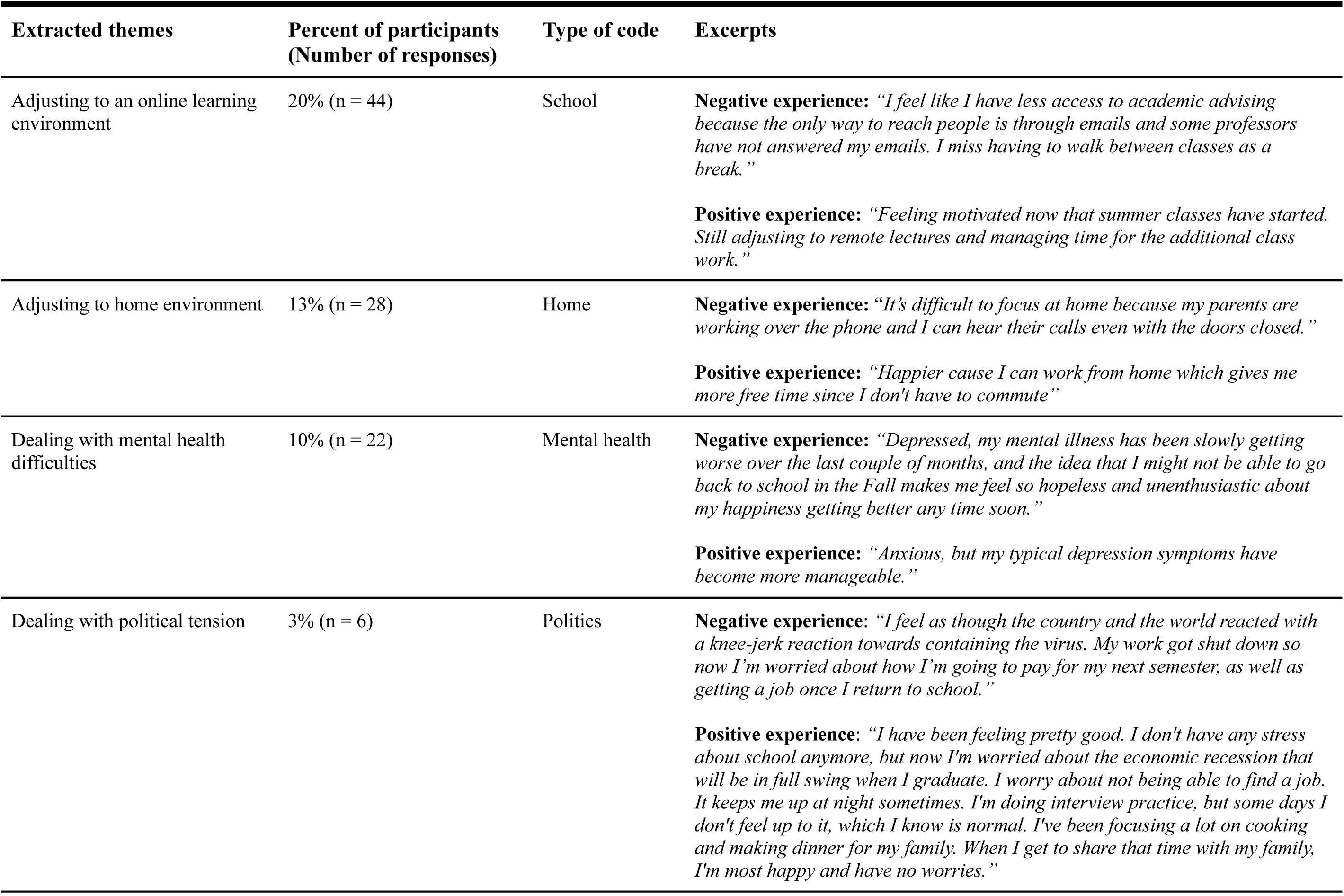
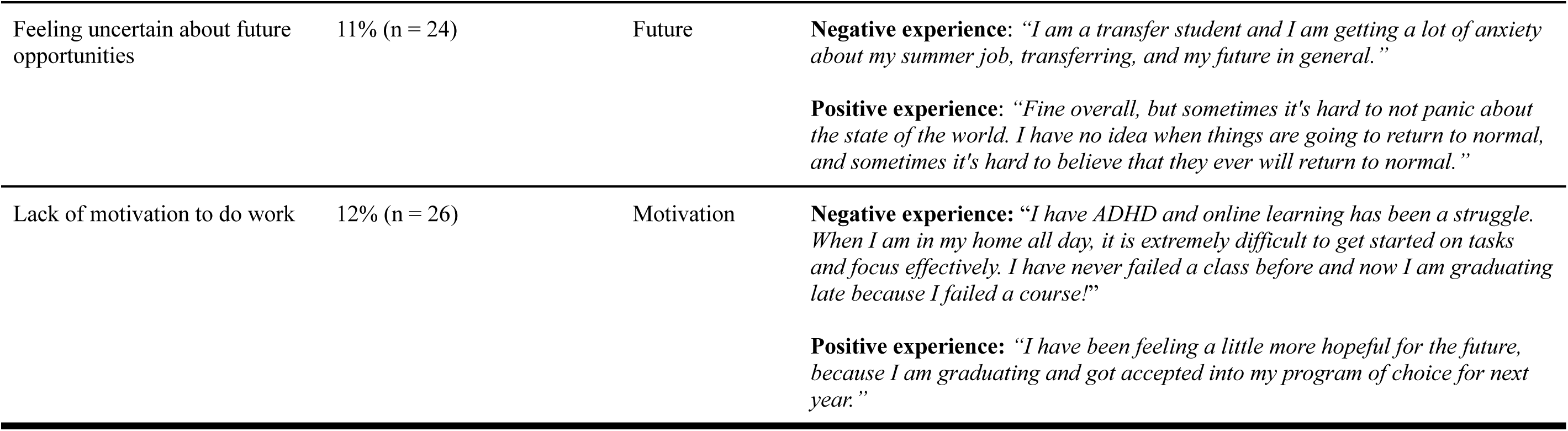
Thematic analysis of open-ended question based on students’ responses (n = 215)

## Discussion

### Summary of results

To our knowledge, this is the first study to explore undergraduate students’ experience during the COVID-19 lockdown between April and June 2020 across over twenty universities in the United States. The results of our study revealed that the majority of participants (n = 802, 68%) reported that moving to remote learning had a negative impact on their academic performance. Overall, we found three significant results in this study: (1) experiencing internet connection issues was one of the predictors of reporting a negative experience with remote learning, (2) students who reported a negative experience with remote learning had higher severity levels of stress, anxiety, and depression, and (3) we identified six common themes associated with positive and negative experiences with remote learning during COVID-19 lockdown.

### Results in context

Students who reported having a negative experience with remote learning may have had more severe and extremely severe levels of stress, anxiety, and depression due to concerns about grades and how that influences access to future opportunities.^19^ While the majority of participants experienced “normal” levels of stress, anxiety, and depression, undergraduate students during lockdown experienced higher levels of “severity” and “extreme severity” compared to other studies conducted prior to the pandemic.^20^ Our results are consistent with articles and reviews that investigated the impact of remote learning on children, adolescents and primary school students.^21,22^ Also, our results regarding remote learning experience are similar to studies conducted outside of the United States.^23,24^

### Limitations

One limitation of this study is that results may not be generalizable to students outside the United States. Lockdown policies may have differed across countries, which may have influenced students’ experience with remote learning during the COVID-19 lockdown. Another limitation is that we excluded all responses from international students, graduate students, and undergraduate students under age 18, which would also impact generalizability to other undergraduate groups in the United States.

### Implications for practice

In light of the findings from this study, future public health research and policies should focus on strategies to better support undergraduate students during future pandemics or similar emergencies that may disrupt traditional learning environments. One important recommendation is for universities and governments to ensure equitable access to reliable internet and technology resources, given that connectivity issues were a significant predictor of negative remote learning experiences. Public health agencies could collaborate with educational institutions to establish contingency plans that provide students with technological support, mental health resources, and academic flexibility during emergencies.

Moreover, addressing mental health should be a key priority in future pandemic preparedness plans. Universities should implement proactive interventions such as virtual mental health counseling, peer support groups, and workshops on coping strategies to mitigate the stress, anxiety, and depression experienced by students. Integrating public health guidelines with educational policies can ensure that mental health support systems are embedded in academic infrastructures.

Additionally, schools should provide students with education and training on how to adapt to studying in the home environment, especially in situations where parents and other family members are present. Workshops or online modules on effective time management, setting boundaries with family, and creating focused study spaces can equip students with practical tools to enhance productivity at home. These strategies are critical to minimizing distractions and stress, fostering a more conducive learning environment, and ensuring students can maintain academic performance during prolonged disruptions.

## Conclusion

Remote learning had a negative impact on the majority of U.S. undergraduate students’ academic performance and mental health during the COVID-19 pandemic. Future research and policy efforts should prioritize strategies to alleviate the negative impacts of sudden educational disruptions during public health crises.

## CRediT authorship contribution statement

**Joseph P. Nano**: conceptualization, methodology, formal analysis, and writing the original draft. **William A. Catterall**: methodology, writing the original draft. **Michael L. Chang:** methodology, writing the original draft. **Mina H. Ghaly:** methodology, formal analysis, writing the original draft.

## Data availability statement

The data which supports the findings of this research are available upon request of the authors.

## Acknowledgments

Special thanks to Dr. Wen Fan from Boston College for all her support over the years.

## APPENDICES

**Appendix 1.**
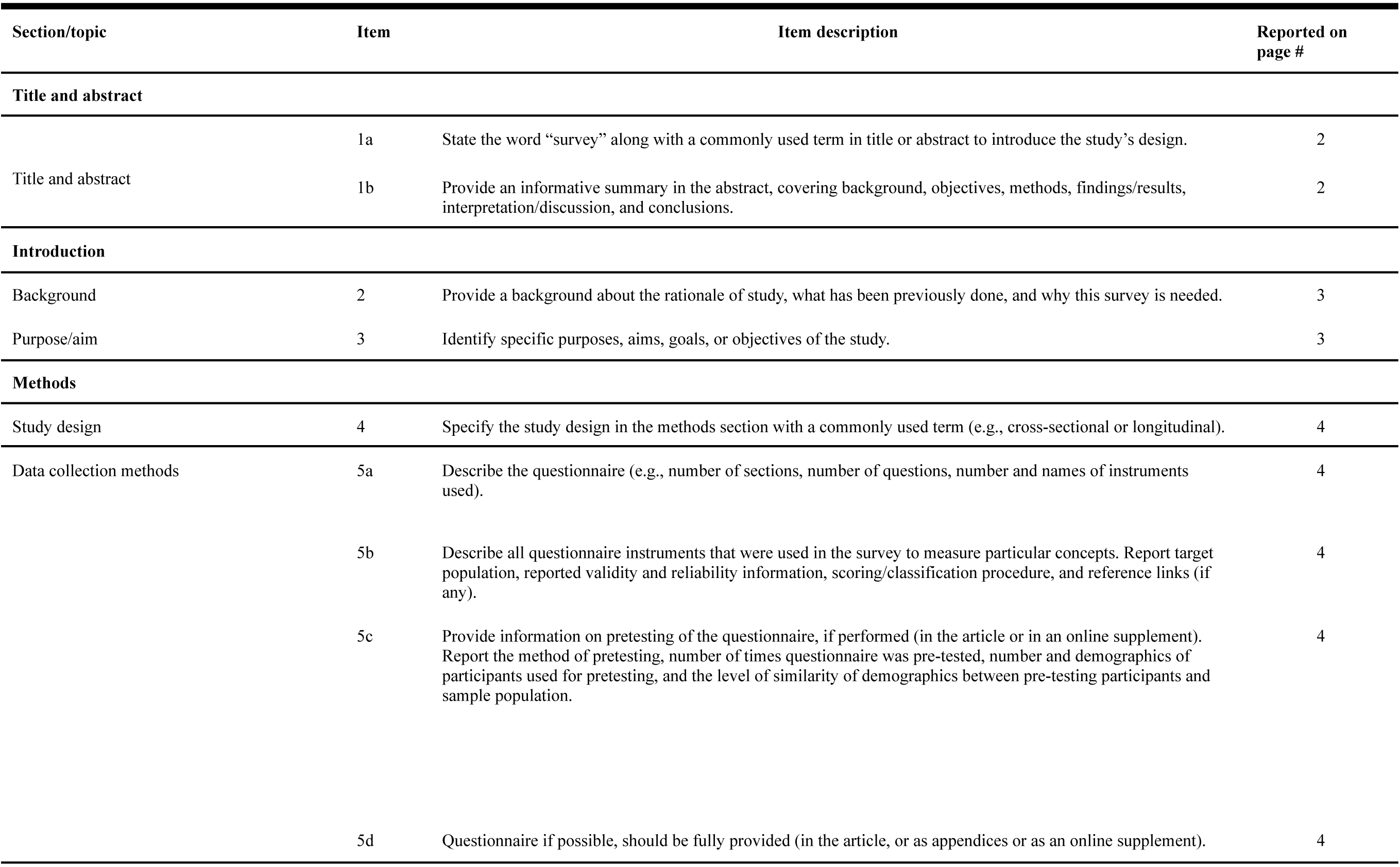
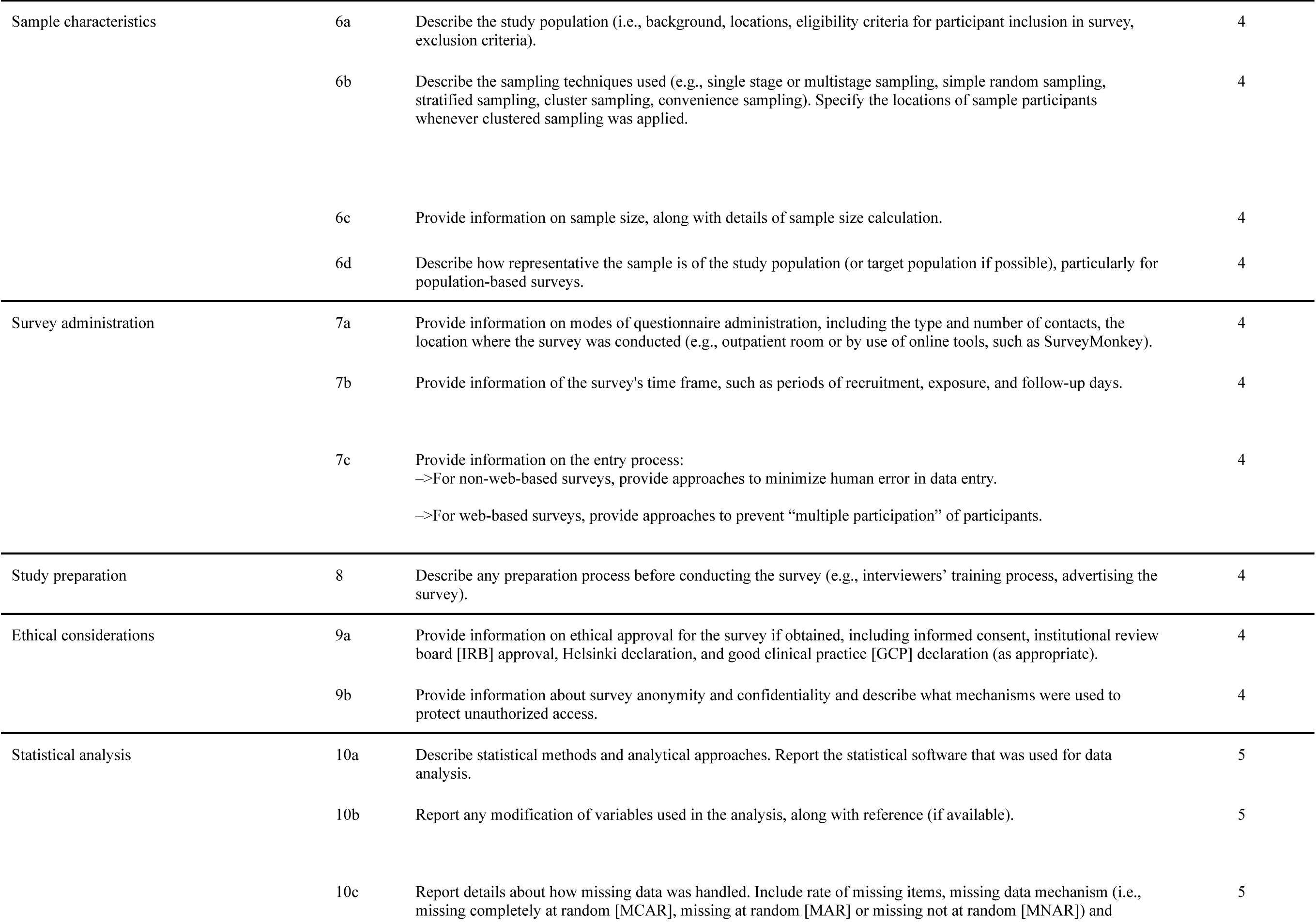
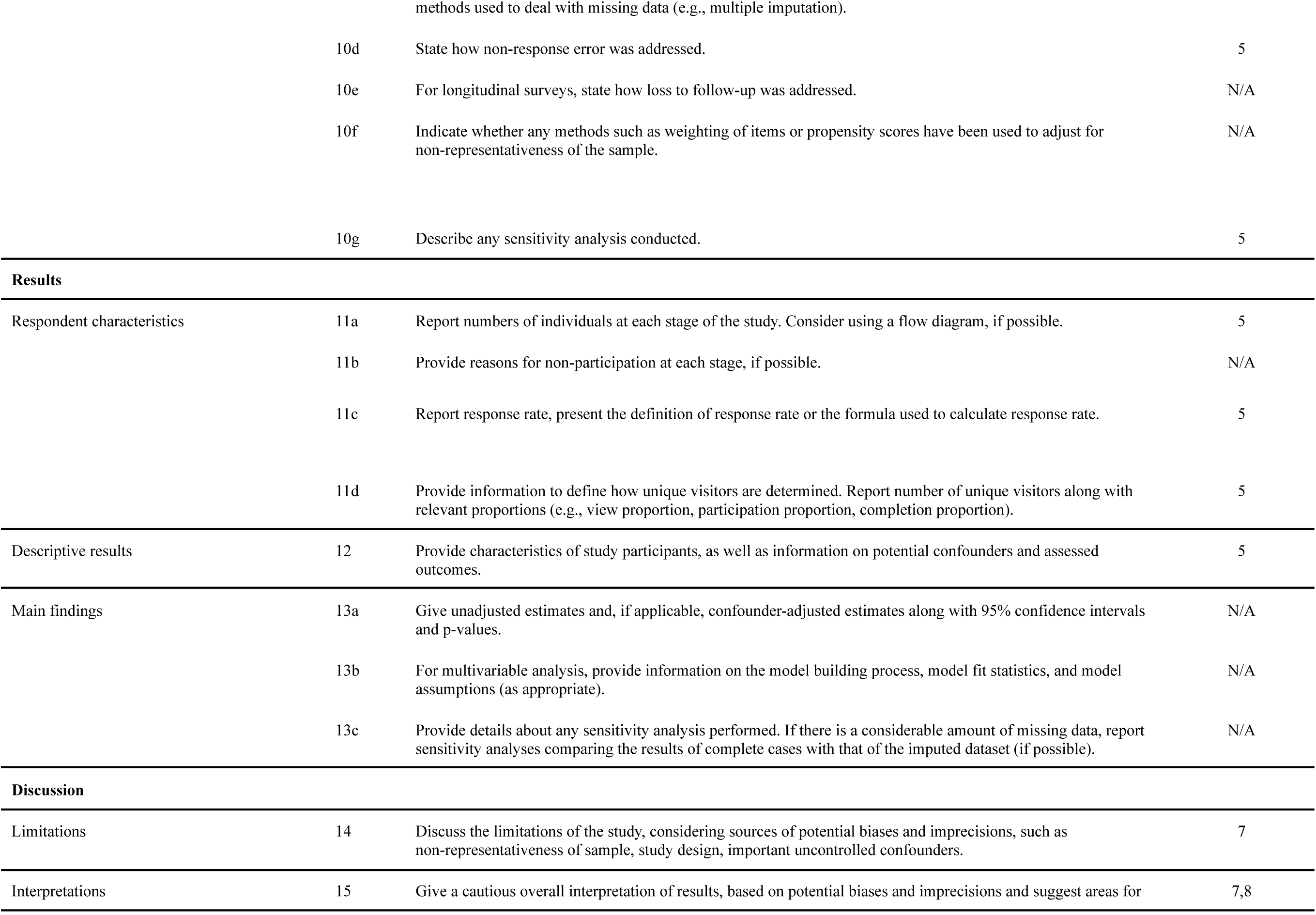
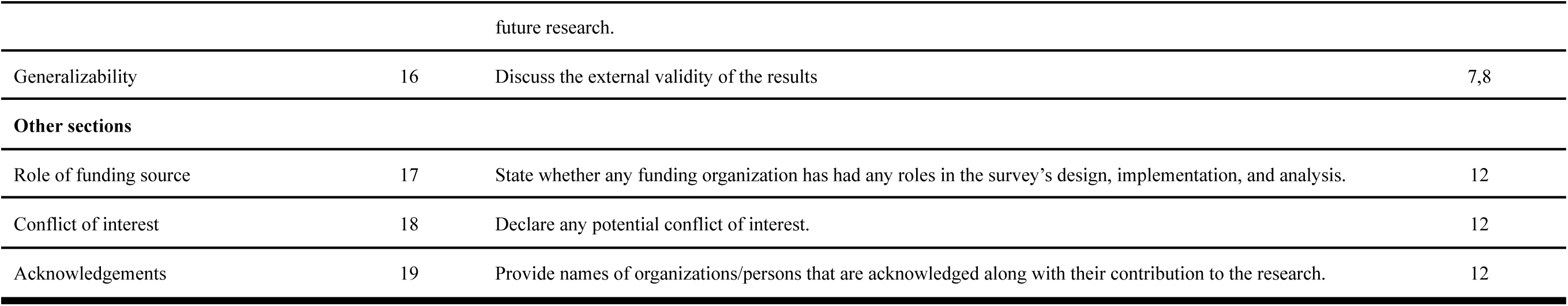
Checklist for Reporting Of Survey Studies (CROSS)^25^.

